# Assessment of ChatGPT in the preclinical management of ophthalmological emergencies – an analysis of ten fictional case vignettes

**DOI:** 10.1101/2023.04.16.23288645

**Authors:** Dominik Knebel, Siegfried Priglinger, Nicolas Scherer, Jakob Siedlecki, Benedikt Schworm

**Affiliations:** Department of Ophthalmology, University Hospital, LMU Munich, Mathildenstraße 8, 80336, Munich, Germany

## Abstract

**Background/Aims:** The artificial intelligence (AI) based platform ChatGPT (Chat Generative Pre-Trained Transformer, OpenAI LP, San Francisco, CA, USA) has gained an impressing popularity over the past months. Its performance on case vignettes of general medical (non-ophthalmological) emergencies has priorly been assessed with very encouraging results. The purpose of this study is to assess the performance of ChatGPT on ophthalmological emergency case vignettes in terms of the main outcome measures triage accuracy, appropriateness of recommended preclinical measures and overall potential to inflict harm to the user/patient.

**Methods:** We wrote ten short, fictional case vignettes describing different acute ophthalmological symptoms. Each vignette was entered into ChatGPT five times with the same wording and following a standardized interaction pathway. The answers were analysed in a standardised manner.

**Results:** We observed a triage accuracy of 87.2%. Most answers contained only appropriate recommendations for preclinical measures. However, an overall potential to inflict harm to users/patients was present in 32% of answers.

**Conclusion:** ChatGPT should not be used as a stand-alone primary source of information about acute ophthalmological symptoms. As AI continues to evolve, its safety and efficacy in the preclinical management of ophthalmological emergencies has to be reassessed regularly.

## Introduction

The artificial-intelligence (AI) platform Chat generative pre-trained transformer (ChatGPT) by OpenAI LP (San Francisco, CA, USA), which is based on the language model generative pre-trained transformer (GPT), constitutes an impressive new tool for generating texts in various contexts and has shown to perform quite well on several academic exams including the Ophthalmic knowledge assessment programme (OKAP). [1, 2] When evaluating the Performance of ChatGPT in medicine, though, one has to keep in mind that it presumably has not been specifically trained on training sets from the medical domain, although the exact training set remains undisclosed.[3] Despite this limitation, the easy accessibility and fast-growing popularity of ChatGPT make it appear likely, that patients may turn to ChatGPT for first information on acute (ophthalmologic) symptoms. Preclinical management and timing of an ophthalmologist’s consultation can largely determine the long-term outcome of ophthalmological emergencies.[4] From a public health perspective, it is therefore crucial to investigate the accuracy and trustworthiness of the information ChatGPT provides in this domain.

ChatGPT has been shown to provide highly accurate general information on retinal disease. [5] However, when it comes to preclinical management of ophthalmological emergencies, three important core tasks have to be mastered: Establishment of an initial set of differential diagnoses and a most probable suspected diagnosis, triaging the patient (i.e. determining the timespan in that he should be referred to an ophthalmologist) and initiation of appropriate preclinical first-aid measures. [4] Encouraging results with regards to differential diagnosis in the general medical domain have been published, [6, 7] and ChatGPT has been found useful for simplifying access to information on cardiopulmonary resuscitation.[8] Despite these encouraging results, ChatGPT can also produce wrong information and has been reported to give potentially harmful advice in the ophthalmological and wider medical domains. [5, 9, 10]

The aim of this study was to evaluate the performance of ChatGPT in the triage and preclinical management of ophthalmological emergencies and to better understand potential chances and risks associated with patients using ChatGPT as a primary source of information about their acute ophthalmological symptoms.

## MATERIALS AND METHODS

### Case vignettes

We created ten case vignettes consisting of simple, short and stereotypic one-sentence descriptions of acute ocular symptoms in English language. They were designed to cover a broad range of ophthalmological subspecialties and to resemble potential patients’ descriptions of acute ophthalmic symptoms. Each vignette was assigned an urgency level as ground truth on the scale “emergency”, “same day”, “same week” and “elective” by authors consensus. The case vignettes and assigned urgency levels are listed in table 1.

**Table 1:** Ten different fictional case descriptions containing short and stereotypic descriptions of acute ophthalmological symptoms were designed to resemble potential patients queries as well as to cover a broad range of ophthalmological subspecialties.

|  | Vignette title | Description text as entered into ChatGPT | Assigned urgency level |
| --- | --- | --- | --- |
| A | Hordeolum | "I have a painful bump on my left upper eyelid." | Elective |
| B | Pediatric leukocoria | "My child has a white pupil." | Same week |
| C | Flashes and floaters | "I see flashes and black spots with my right eye." | Same day |
| D | Sudden monocular vision loss | "I suddenly can't see with my left eye." | Emergency |
| E | Sudden, painful monocular vision loss | "My right eye suddenly hurts, and I can't see any more." | Emergency |
| F | Sudden onset diplopia | "All of a sudden, I see everything double." | Emergency |
| G | Dry eye | "My eyes burn and itch." | Elective |
| H | Monocular red eye | "My right eye is red since yesterday." | Same week if persistent mild to moderate symptoms, same day if severe |
| I | Corneal erosion | "My toddler scratched my right eye, now it hurts and is red." | Same day |
| J | Alkali burn | "My colleague got mortar dust into his left eye, now he can't open it because it hurts so much." | Emergency |

### Standardized pathway for interaction with ChatGPT

We used the free research preview of ChatGPT in the version of March 14, 2023. We entered the case description followed by a question for diagnosis and treatment recommendation (“question 1”) into ChatGPT. Depending on whether or not the answer generated by ChatGPT (“answer 1”) contained the unconditional recommendation to visit a physician, we entered one of two different second questions into ChatGPT (“question 2”). If upon viewing the answer generated by ChatGPT (“answer 2”) in combination with answer 1 we felt any need for further inquiry or clarification, an optional, non-standardized third question (“question 3”) would be allowed. The complete standardized interaction pathway is depicted in diagram 1. For each case vignette, this standardized pathway was repeated five times with the same wording. The repetition of queries has been priorly used by other study groups, because ChatGPT generates a new answer at each attempt, with answers potentially differing from instance to instance. [5, 9]

### Analysis of answers generated by ChatGPT

The 50 responses generated by ChatGPT were analysed in a systematic approach, with the individual responses to the first and second questions of the standardized pathway being analysed separately and together. The detailed manual for evaluation of the answers can be found in the supplements. The main outcome measures of our analysis were triage accuracy on global and vignette levels and appropriateness of recommended preclinical measures as well as overall potential of inflicting harm to the user/patient on the level of individual attempts. Triage accuracy was defined as the share of attempts, for which the urgency level stated in answer 2 matched the ground truth, i.e., the urgency level assigned to the vignette by the authors. The appropriateness of the preclinical measures (APM) recommended in answer 2 was graded on a five-point ordinal scale as indicated in table 2. For each attempt, answers 1, 2 and 3 combined were judged with regards to their overall potential to inflict harm on a binary scale (yes/no). All other investigated parameters were evaluated as described in the supplements and constitute secondary outcome measures. A descriptive statistical analysis was performed using Microsoft Excel (Microsoft Corporation, Redmond, United States).

**Table 2:**
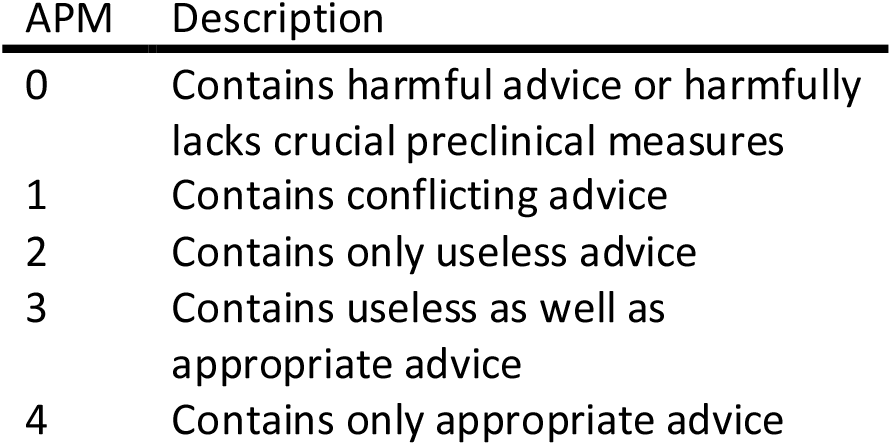
Appropriateness of recommended preclinical measures (APM) was graded on a five-point ordinal scale.

## RESULTS

### Unconditional recommendation to consult a physician

In 3 of 50 attempts (6%), all concerning case vignette A (“Hordeolum”), no unconditional recommendation to consult a physician was contained in answer 1. In 2 of those 3 attempts, even question 2 “Are you sure, or should I/we rather see a doctor right away?” did not lead ChatGPT to integrate such a recommendation in answer 2. In the standardized interaction pathway, the remaining majority of 47 attempts (94%) (with answer 1 containing that recommendation) led to question 2 asking about urgency and preclinical measures, hence only those are analysed with regards to triage and preclinical measures.

### Diagnosis

Answer 1 contained a single diagnosis in 12 of 50 attempts (24%) and a most probable diagnosis among a list of differential diagnoses in one attempt (2%). In the majority of attempts (32 of 50 attempts, 64%), answer 1 contained a list of differential diagnoses without specifying a most probable diagnosis. In five attempts, answer 1 contained no diagnosis at all.

Overall diagnostic accuracy was 61.5% in the attempts where a single most probable diagnosis was given (8 of 13 correct).

### Treatment

Specific advice (i.e. “treatment for your condition is A”) was given in answer 1 in 10 of 50 instances (20%). General information (i.e. “treatment for condition 1 would be A, for condition 2 would be B, …”) with regards to treatment was contained in 10 instances (20%). In the remaining 30 instances (60%), only vague or no information on treatment were given. Overall treatment accuracy was 100%, that is if specific treatment advice and a single most probable diagnosis were given, treatment advice would have been appropriate for this diagnosis in 8 of 8 attempts (100%), regardless of correctness of the given diagnosis.

### Triage

Information on urgency were contained in 47 of 47 instances (100%). Overall triage accuracy was 87.2%, that is answer 2 contained a reasonable advice on urgency in 41 of the 47 attempts (87.2%). Urgency was overestimated in 6 attempts (12.8%) and underestimated in 0 attempts (0%). In one instance, question 3 was asked to clarify the urgency level.

### Preclinical measures

Answer 2 contained recommendations for preclinical measures in 47 of the 47 attempts where question 2 asked for them (100%). Overall median APM was 4 (only appropriate measures), ranging from 0 (harmful advice) to 4 (only appropriate measures). Answer 2 contained only appropriate measures in 31 attempts (66.0%), appropriate measures as well as useless ones in 7 attempts (14.9%), only useless measures in 2 attempts (4.3%) and conflicting advice in one attempt (2.1%). In 4 attempts (8.5%), answer 2 contained potentially harmful advice such as patching the unaffected fellow eye in pediatric leukocoria before establishing a proper diagnosis (note that the age of the child was not specified in case description B). In 2 attempts (4.3%), the advice was potentially harmful because it lacked or largely understated the crucial recommendation to immediately irrigate the eye affected by a suspected alkali burn.

### Overall evaluation of answers 1, 2 and 3

The answers contained questions directed at the user in 0 of 50 instances (0%). Wrong information was contained in 12 instances (24%) and conflicting advice in 18 instances (36%). Wrong information came frequently in form of wrong differential diagnoses (for example retinal detachment as differential diagnosis for sudden painful monocular vision loss) or misconceptions about preclinical measures, such as the misconception that in sudden monocular vision loss patching the unaffected fellow eye may improve the vision of the affected eye. Conflicting advice was frequently given with regards to the urgency, i.e. the appropriate timespan in which to consult a physician. Overall, the severity of the symptoms was captured correctly in 38 instances (76%), rather overestimated in 5 instances (10%) and rather underestimated in 7 instances (14%). Overall, 16 responses (32%) were judged to carry the potential to inflict harm to a patient following the contained recommendations.

### Explicit disclaimer and vignette-level results

Some form of explicit disclaimer that ChatGPT cannot provide a diagnosis or medical advice was contained in 31 of 50 instances (62%). Table 3 sums up the performance of ChatGPT on the individual vignettes. On a vignette level, there was little linear correlation between the number of answers per vignette that contained such a disclaimer on the one hand, and the vignette-level rate of harmful answers and vignette-level diagnostic accuracy on the other hand, as visualized in figure 2. A Spearman’s rank correlation coefficient of -0.15 shows a weak negative correlation between the number of answers per vignette containing a disclaimer and vignette-level median APM, which would be expected if the presence of a disclaimer indeed indicated less appropriate recommended preclinical measures. In contrast, a Spearman’s rank correlation coefficient of 0.13 indicates a weak positive correlation between the number of answers per vignette containing a disclaimer and vignette-level minimum APM.

**Table 3:** Performance of ChatGPT on the individual vignettes. APM = Appropriateness of recommended preclinical measures, graded on the five-point ordinal scale presented in table 2, where 0 indicates a harmful recommendation and 4 indicates a completely appropriate recommendation.

|  | Vignette title | Diagnostic accuracy | Disclaimer contained | Triage Accuracy | APM [median, [range]] | Potentially harmful answers |
| --- | --- | --- | --- | --- | --- | --- |
| A | Hordeolum | 5/5 (100%) | 0/5 (0%) | 2/2 (100%) | 4, [4; 4] | 0/5 (0%) |
| B | Pediatric leukocoria | 0/1 (0%) | 2/5 (40%) | 1/5 (20%) | 3, [0; 4] | 1/5 (20%) |
| C | Flashes and floaters | 0/1 (0%) | 5/5 (100%) | 5/5 (100%) | 3, [3; 3] | 0/5 (0%) |
| D | Sudden monocular vision loss | - | 5/5 (100%) | 5/5 (100%) | 1, [0; 3] | 4/5 (80%) |
| E | Sudden, painful monocular vision loss | - | 5/5 (100%) | 5/5 (100%) | 4, [4; 4] | 0/5 (0%) |
| F | Sudden onset diplopia | - | 0/5 (0%) | 5/5 (100%) | 4, [4; 4] | 0/5 (0%) |
| G | Dry eye | - | 5/5 (100%) | 4/5 (80%) | 4, [4; 4] | 0/5 (0%) |
| H | Monocular red eye | - | 5/5 (100%) | 4/5 (80%) | 4, [4; 4] | 5/5 (100%) |
| I | Corneal erosion | 3/3 (100%) | 3/5 (60%) | 5/5 (100%) | 4, [0; 4] | 1/5 (20%) |
| J | Alkali burn | 0/3 (0%) | 0/5 (0%) | 5/5 (100%) | 4, [0; 4] | 5/5 (100%) |

**Figure 1:**
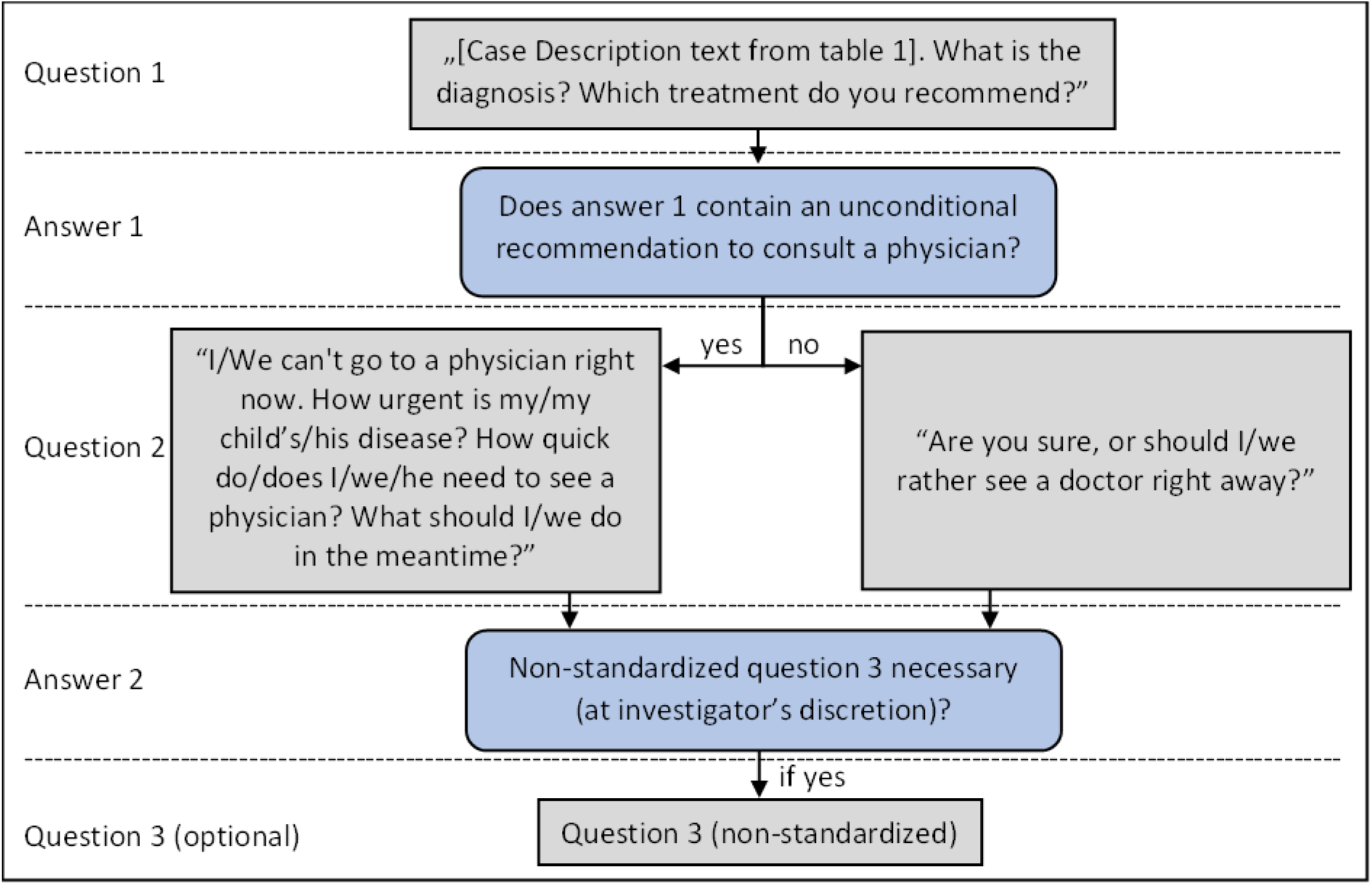
Data was generated via interaction with ChatGPT following a standardized pathway, on which two or three questions were entered into ChatGPT sequentially, with question 2 and 3 depending on the answers generated by ChatGPT.

**Figure 2:**
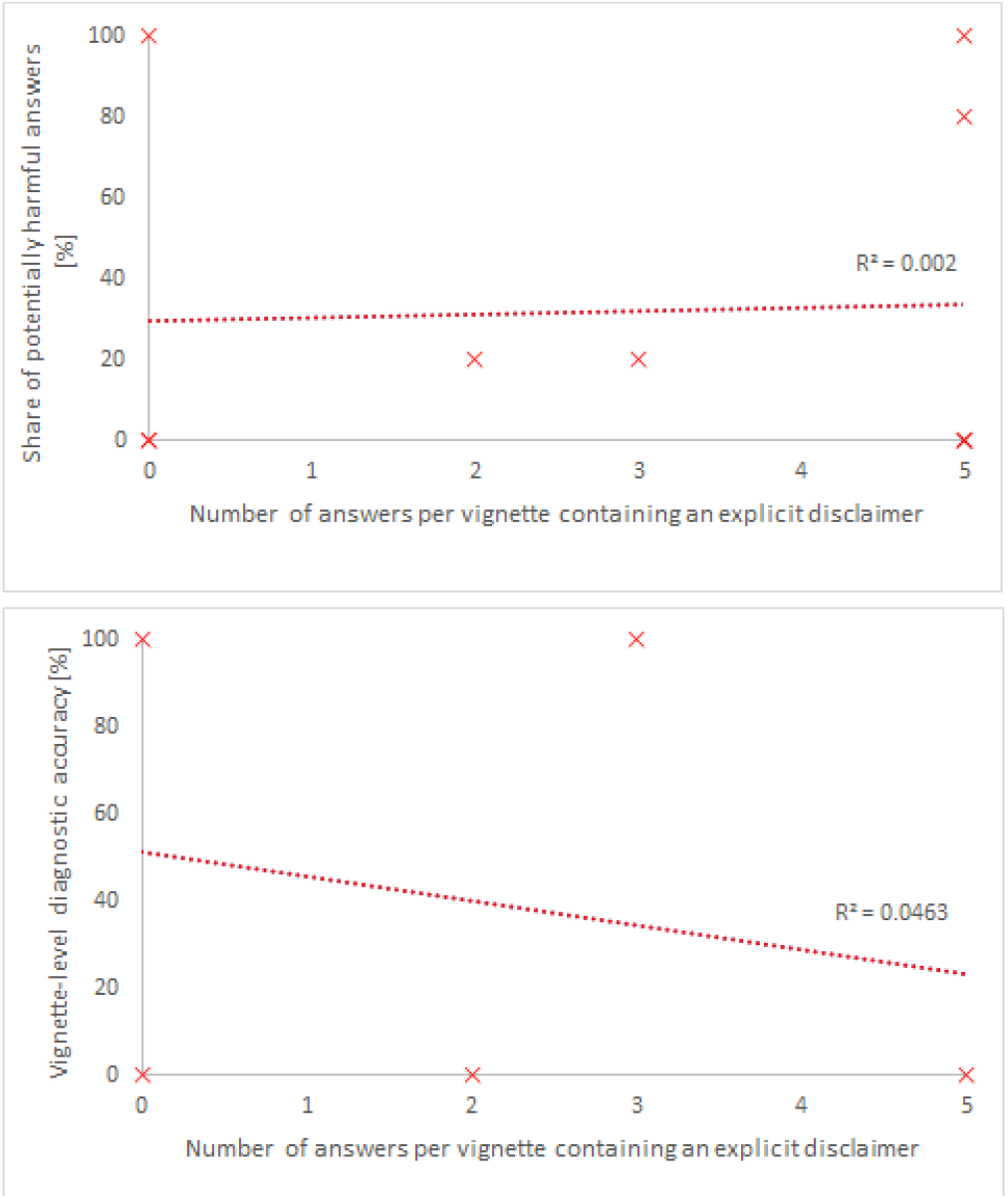
The number of answers per vignette that contain an explicit disclaimer warning that ChatGPT can’t give a diagnosis or medical advice shows weak correlations with vignette-level accuracy measures. Diagnostic accuracy tends to by lower, when the answers for a vignette contained such a disclaimer more often. This would be expected, if the presence of the disclaimer indeed indicates a lower certainty of the answer. The vignette-level share of potentially harmful answers shows nearly no correlation to the number of answers containing a disclaimer.

## DISCUSSION

At first glance, ChatGPT performs remarkably well in terms of accuracy for treatment (100%), triage (87.2%) and diagnosis (61.5%) of ophthalmological emergencies as well as the appropriateness of recommended preclinical measures (APM, overall median 4), especially given that it has not been exposed to specific training sets from the ophthalmological or even from the general medical domain. However, following the recommendations of ChatGPT would potentially lead to harm in 32% of the investigated conversations.

A very encouraging triage accuracy of 87.2% in our study stands in contrast to recent results on the non-ophthalmological general medical domain published in a preprint by Levine and colleagues, who found a triage accuracy of 71% for GPT-3 and 96% for physicians. [7] Whether this contrast is due to the different testing domains, wording of the individual vignettes or an improvement between the different GPT versions remains unclear. Moreover, we must point out, that a technically high triage accuracy does not imply a great utility of the information on urgency: In our study ChatGPT frequently recommended to consult a physician “as soon as possible”, which was judged to be appropriate for the urgency levels “emergency” and “same day”. The technically high triage accuracy therefore came with the downside of a certain vagueness. While technically being appropriate in the vast majority of cases, the lack of nuance to distinguish between those two urgency levels might lead a patient with a suspected corneal erosion to rather overestimate the urgency of his condition, whereas the need of immediate medical attention for a patient with sudden onset diplopia might be understated by the answers generated by ChatGPT.

A similar pattern evolves with regards to treatment accuracy: While a perfect treatment accuracy (100%) was reached in those instances where specific treatment advice and a single most probable diagnosis were given, in 80% of all instances ChatGPT gave only vague or general treatment information if any. In contrast to our observed treatment accuracy, Potapenko et al. reported less accuracy of ChatGPT in terms of treatment than in terms of diagnosis, prognosis or general information concerning retinal diseases. [5]

Furthermore, it is noteworthy that our vignettes were not designed to measure treatment or diagnostic accuracy, but rather to resemble potential queries of emergency patients. Therefore, many of them do not provide enough information to narrow down the list of differential diagnoses to one single diagnosis and give specific treatment advice. We also did not ask ChatGPT to provide a list of differential diagnoses or to elaborate on treatment options for these. It is therefore actually quite impressive, that ChatGPT provided such lists in cases where it could not give a single diagnosis. In our analysis of diagnostic accuracy, though, we included only those answers that did specify a single (most probable) diagnosis. Our study design might therefore explain why the observed diagnostic accuracy (61.5%) was lower than the observed treatment and triage accuracies. In comparison, Levine et al. and Hirosawa et al. report much higher diagnostic accuracies (88% and 93.3%, respectively) on fictional case vignettes of non-ophthalmological emergencies. This might be explained by ophthalmology being a more specialised domain than general medicine, therefore possibly being less strongly represented in the training sets of GPT, which have not yet been disclosed to the public. [3] Moreover, as ophthalmology is a specialty which heavily relies on visualization of the ocular structures to establish a diagnosis, the possibility to establish diagnosis based on verbal patient statements might be generally limited, for ChatGPT as well as for ophthalmologists. Indeed, a study from the Wills Eye emergency department found the diagnostic accuracy of ophthalmologists triaging via telephone to be 69.9%,[11] only slightly above the diagnostic accuracy of ChatGPT we observed.

However, analysis of diagnostic accuracy on vignette level (see table 3) showed only values of 0% or 100%. This can be very problematic in an actual ophthalmological emergency, as ChatGPT might produce very accurate answers in some cases and very inaccurate answers in others. To the emergency patient, it remains unclear whether his case is one of the accurately answered or one of the inaccurately answered ones. Furthermore, ChatGPT does not provide any information on the sources its answers are based on. However, newer versions of ChatGPT integrated into Microsoft’s Bing engine have been updated to integrate references on its sources. [12]

The advice on preclinical measures given by ChatGPT was appropriate in the majority of attempts (66%), hence median APM was 4. However, ChatGPT gave potentially harmful advice in 12.7% of attempts. This very closely matches the results published by Potapenko et al., who identified harmful treatment advice in 12 of 100 answers with regards to retinal diseases. [5]

At least, the answers in our analysis frequently contained an explicit disclaimer warning that ChatGPT cannot give a diagnosis or medical advice. However, the frequency at which this disclaimer was produced per vignette did only weakly correlate with important vignette-level accuracy measures (see figure 2), and the absence of such a disclaimer did not indicate the absence of any potential to inflict harm to the user/patient.

In our analysis, we observed an overall potential to inflict harm in 32% of the answers. For now, we therefore clearly recommend not to use ChatGPT as primary source of information on acute ocular symptoms. Yet it remains unclear, if non-professionals (with or without the possibility to obtain information through the internet) perform better or worse than ChatGPT in the management of ophthalmological emergencies. In the domain of general medical emergencies, Levine et al. found the diagnostic accuracy of GPT-3 to be significantly superior to lay persons, indicating that they might profit from the use of GPT-3. Triage accuracy however was slightly and insignificantly lower compared to laypersons but by far and significantly lower compared to physicians. [7] For “can’t-miss diagnoses”, the aforementioned study from the Wills Eye emergency department showed a diagnostic accuracy of triaging ophthalmology staff to be as high as 97.2%. [11] We therefore clearly recommend contacting established providers of ophthalmological emergency services in case of acute symptoms.

A limitation to our study is, that we used carefully worded case vignettes instead of real patient queries. Those may differ in many ways from our vignettes, for example potentially containing more vague statements or confounding and conflicting information, and therefore ChatGPT may react differently to them.

A study from Teebagy et al. that was recently published as preprint has compared the versions of ChatGPT accessible in March 2023 and December 2022 with regard to performance on the OKAP exam. Their study shows a remarkable and significant improvement from 57% correct answers in December to 81% correct answers in March. This fast and impressive improvement of a general AI-based language model in the ophthalmological domain and the potential of language models specifically pre-trained in the medical domain are very encouraging.

Although ChatGPT should presently not be consulted for acute ocular symptoms, it already shows very impressive capabilities in the ophthalmological domain. As AI based language models continue to improve, we believe that they will soon start to play a more important role in the preclinical management of ophthalmological emergencies. We should embrace those technologies and continue to seek for a better understanding of the strengths and limitations of AI-based language models in the context of clinical ophthalmology.

## Data Availability

All data produced in the present study are available upon reasonable request to the authors

---

**Supplementary Table 1:**
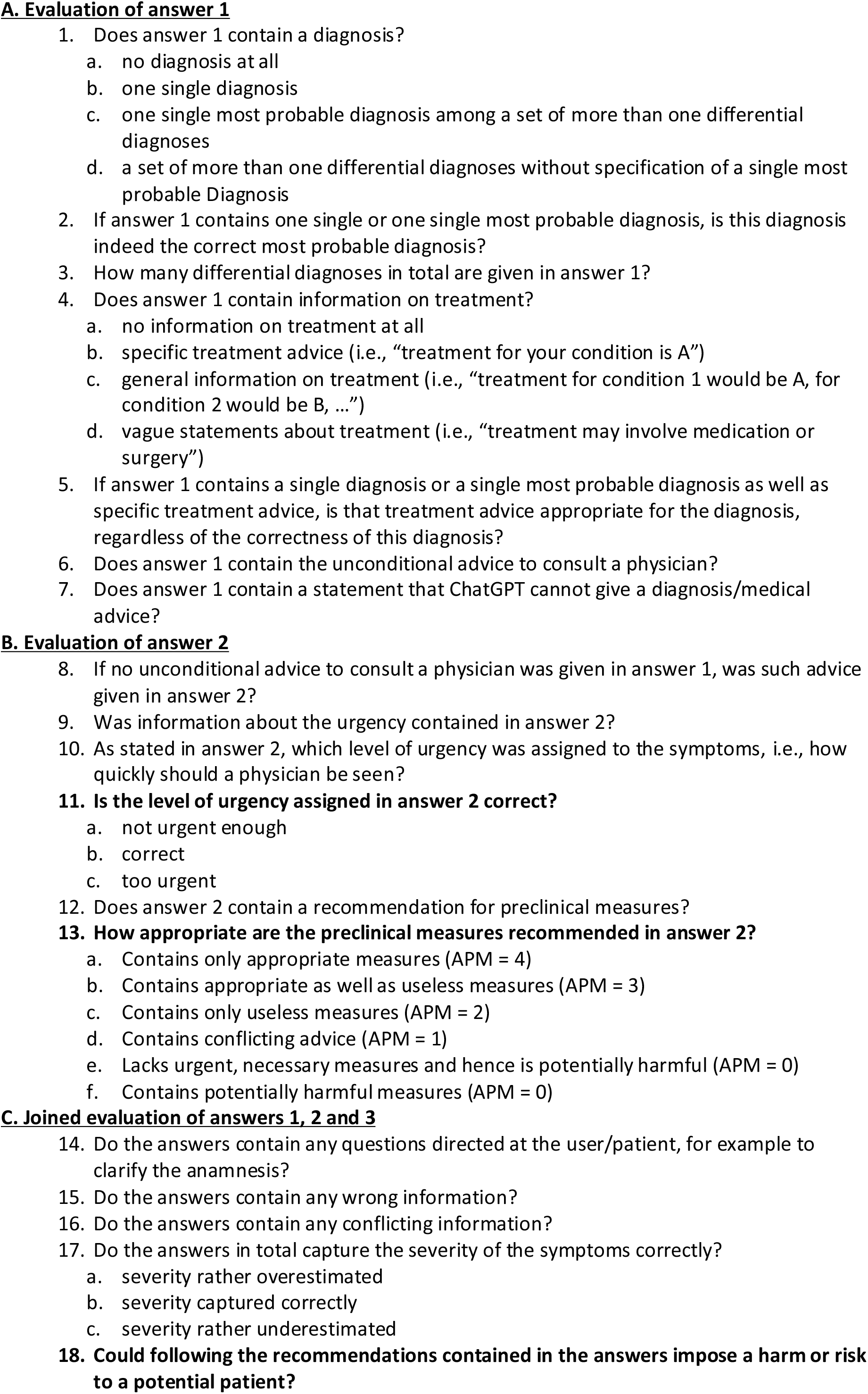
Detailed Manual for evaluation of the answers generated by ChatGPT. The answers 1, 2 and 3 generated by ChatGPT were analysed in a structured manner with regards to 18 different aspects. Variables relevant to main outcome measures are marked in bold. APM: Appropriateness of preclinical measures

